# Selective ensemble methods for deep learning segmentation of major vessels in invasive coronary angiography

**DOI:** 10.1101/2021.09.13.21263481

**Authors:** Jeeone Park, Jihoon Kweon, Hyehyeon Bark, Young In Kim, Inwook Back, Jihye Chae, Jae-Hyung Roh, Do-Yoon Kang, Pil Hyung Lee, Jung-Min Ahn, Soo-Jin Kang, Duk-Woo Park, Seung-Whan Lee, Cheol Whan Lee, Seong-Wook Park, Seung-Jung Park, Young-Hak Kim

## Abstract

Invasive coronary angiography is a primary imaging modality that visualizes the lumen area of coronary arteries for the diagnosis of coronary artery diseases and guidance for interventional devices. In the current practice of quantitative coronary analysis (QCA), semi-automatic segmentation tools require labor-intensive and time-consuming manual correction; this limits their application in the catheterization room. For a more automated QCA, it is necessary to minimize operator intervention through robust segmentation methods with improved predictability. In this study, we introduced two selective ensemble methods that integrated the weighted ensemble approach with per-image quality estimation. In our selective ensemble methods, the segmentation outcomes from five base models with different loss functions were ranked by mask morphology or estimated dice similarity coefficient (DSC). The final output was determined by imposing different weights according to the ranking. The ranking criteria based on mask morphology were determined empirically to avoid frequent types of segmentation errors, whereas the estimation of DSCs was performed by comparing the pseudo-ground truth generated from a meta-learner. In the assessment with 7,426 frames from 2,924 patients, the selective ensemble methods improved segmentation performance with DSCs of up to 93.11% and provided a better delineation of lumen boundaries near the coronary lesion with local DSCs of up to 94.04%, outperforming all individual models and hard voting ensembles. The probability of mask disconnection at the most narrowed region could be minimized to <1%. The robustness of the proposed methods was evident in the external validation. Inference time for major vessel segmentation was approximately one-third, indicating that our selective ensemble methods may allow the real-time application of QCA-based diagnostic methods in routine clinical settings.

## 1 Introduction

Cardiovascular diseases (CVDs) are the most prevalent cause of death worldwide, representing 32.2% of the total mortality; 49.7% of CVD-associated mortalities are coronary artery diseases (CADs) (World Health Organization, 2019). Obstruction of the coronary artery causes the restriction of blood flow to the heart muscle, a condition known as myocardial ischemia. For the revascularization of obstructed vessels, percutaneous coronary intervention with balloon angioplasty and stent implantation is the standard treatment.

Invasive coronary angiography (ICA) is a primary imaging modality that visualizes the lumen area of coronary arteries for the diagnosis of CAD and guidance for interventional devices. Through the delineation of vessel boundaries, quantitative coronary analysis (QCA) provides objective parameters for clinical decisions. Using QCA, Synergy between Percutaneous Coronary Intervention with Taxus and Cardiac Surgery (SYNTAX) scores allow a comprehensive assessment of coronary arteries with multi-vessel disease (Sianos et al., 2005; Thuijs et al., 2019). The functional evaluation of stenosed lesions (Morris et al., 2020; Tu et al., 2014) and the detection of unstable atherosclerotic plaque (Stone et al., 2012; Tomaniak et al., 2020) could be performed with a three-dimensional (3D) reconstruction of coronary geometry through the integration of QCA from multiple views.

In the current practice, QCA is performed by manually correcting the initial mask acquired using semi-automatic edge-detection tools. Due to the anatomical diversity of coronary arteries and imaging artifacts (Blondel et al., 2006), considerable experience is required to accurately perform QCA. Manual corrections could prolong the duration of the interventional procedure. Although various automated methods using image processing techniques have been proposed to mitigate the workload for QCA (Cruz-Aceves et al., 2016; Fazlali et al., 2018; Qin et al., 2019), they have not been adopted in the routine clinical setting due to their prediction accuracy and processing time.

Recent advances in segmentation methods using deep networks have demonstrated the improved predictability of the vessel areas in ICA (Iyer et al., 2021; Jun et al., 2020; Yang et al., 2019). Vessel overlaps, which hinder the accurate identification of coronary arteries, were alleviated with multi-view learning (Zhang et al., 2020). In addition, the coronary lesions could be localized without the direct segmentation of the luminal area (Moon et al., 2021). Nonetheless, images that yield a low segmentation performance from deep learning models require more manual correction compared to the conventional tool using the image processing method (Yang et al., 2019). Considering the real-time application in the catheterization room, it is necessary to minimize serious segmentation errors that require the operator’s intervention.

### 1.1 Related works

The application of deep learning has considerably improved the ICA segmentation performance (Silva et al., 2021; Zhao et al., 2021; Zhu et al., 2021). Yang et al. (2019) evaluated the segmentation performance for three major vessels using a large database of ICA images and U-Net architecture, exhibiting a dice similarity coefficient (DSC) of 91.7%. Jun et al. (2020) proposed a nested encoder-decoder architecture for ICA images. Iyer et al. (2021) proposed an integration of segmentation architecture with a preprocessing network as the trained filters for boundary sharpening and contrast enhancement of ICA. Real-time segmentation to capture the dynamic motion of coronary arteries could be performed using one-shot video object segmentation (Zhao et al., 2018). Recently, multi-view approaches using ICA sequences have been actively conducted (Liang et al., 2020; Wan et al., 2021; Wang et al., 2020). In a study by Zhang et al. (2020), discriminative information on the ICA images acquired from different projection angles was trained by adopting intra-view hierarchical attentive blocks.

The fundamentals for improving the prediction performance using the ensemble method is the diversity in base classifiers (Brown et al., 2005; Dietterich, 2000). Over the past decades, numerous approaches have been proposed to enhance and quantify the ensemble diversity (Ganaie et al., 2021). Recent ensemble studies for medical imaging achieved a remarkable improvement with multi-view (Sundaresan et al., 2021), various preprocessing (Lyu et al., 2020), and network architecture (Sohail et al., 2021). Neural architecture search (NAS) was also used to optimize the composition of the base classifiers for the ensemble approach (Baldeon Calisto and Lai-Yuen, 2020). For the prediction diversity, the featured loss functions could be applied. For the segmentation of medical imaging, many attempts have been made to improve the performance with a novel loss function suitable for the problem to be solved (Byrne et al., 2021; Caliva et al., 2019; Gerl et al., 2020; EL Jurdi et al., 2021). In particular, in highly imbalanced problems, the incorporation of a regional loss with the auxiliary loss to evaluate the morphological features of predicted masks was shown to augment the predictive capability (Kervadec et al., 2021; Shit et al., 2021).

Determining the optimal method to combine base classifiers is critical to the performance of the final outcomes from ensemble models (Zhou et al., 2002). The most widely used combination rule is naive averaging, which has demonstrated significant improvement in predictive accuracy for the semantic segmentation of medical imaging (Couteaux et al., 2019; Kang and Gwak, 2019; Murugesan et al., 2021; Weisman et al., 2020; Winzeck et al., 2019). Chassagnon et al. (2020) proposed AtlasNet using an ensemble of six convolutional neural networks to evaluate the extent of interstitial lung disease in systemic sclerosis; this showed a comparable performance to that of radiologists. The weight of each classifier can be determined proportionally with regard to the average performance in a validation set, such as accuracy, area under the curve, and DSC (Mondal et al., 2021). Zhai et al. (2020) proposed an ensemble method that imposed different weights to myocardium edema and scars in the aggregation of two-dimensional (2D) and 2.5D magnetic resonance imaging (MRI). The stacking scheme is a generalized form of the weighted averaging. Zheng et al. (2019) proposed a fully convolutional network-based meta-learner that is trained using the outputs of 3D and 2D base-learners.

Estimating the prediction quality of individual segmentation is another way to improve the performance using multiple outcomes. Recently, Hann et al. (2021) proposed a quality control-based framework for the segmentation of cardiac MRI T1 mapping that pick an output estimated to have the highest DSC among single and label-voting models. The DSC estimation was achieved using the DSC association matrix that correlated the final outcome with the DSC value of each model, which was constructed by applying linear regression to the training data.

### 1.2 Contributions

In this research, we proposed selective ensemble methods that integrated the weighted ensemble approach with per-image quality estimation for the ICA segmentation. The key contributions of our study are as follows:

1. We proposed two ranking rules to assess the major vessel segmentations from prediction models, which were determined using mask morphology and estimated DSC using meta-learners.
2. We adopted novel loss functions to employ the diversity of prediction outcomes. The loss functions were carefully tailored to tackle the error patterns frequently seen in previous studies.
3. We evaluated the performance of deep learning models for ICA segmentation using a database containing 7,426 labeled ICA images acquired from what we know to be the largest number of patients to date and conducted external validation. All of the patients for whom QCA was applied in routine clinical practice were included.
4. For the practical application of automated QCA, we analyzed the evaluation metrics in the local window near the coronary lesion and performed the major blob analysis. Severe prediction errors that hindered automated QCA were classified by ICA image.

## 2 Materials and Methods

In this study, we proposed two selective ensemble methods that produced a better prediction of major vessel areas for automated QCA. Both selective ensemble methods comprised 1) obtaining prediction results from segmentation models focusing on different morphological features, 2) ranking the prediction results based on morphological characteristics or estimated DSC, and 3) combining the prediction results with weights that varied according to the ranking (Fig. 1). For the major vessel segmentation in ICA, an integration of U-Net with DenseNet-121 Huang et al. (2017) was applied, and five different loss functions provided diversity in the prediction outcomes (Fig. 1a).

**Figure 1:**
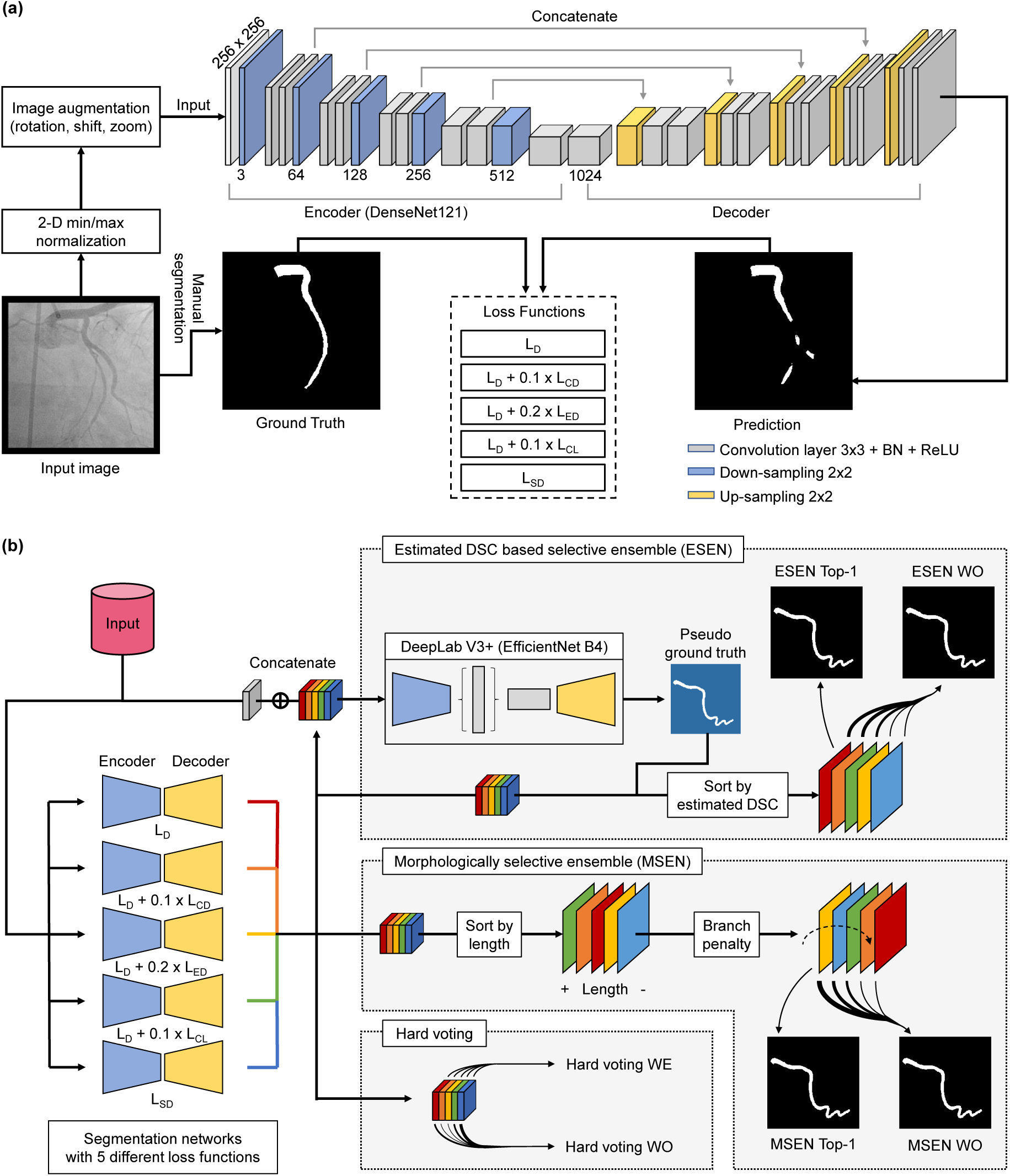
Overview of selective ensemble method using per-image quality estimation (a) Schematic diagram of deep learning approaches. The major vessel areas (marked white) in the predicted mask were evaluated with five different loss functions. (b) Application process of ensemble models using the outputs from five individual models. The hard voting models imposed fixed weights to the individual models (colored boxes). In our selective ensemble models, the predictions were weighted differently for individual images (colored rectangles) according to the ranking. The ranking was determined by morphology and estimated DSC for MSEN and ESEN models, respectively.

### 2.1 Loss functions

Let *x* : Ω ⊂ ℝ^2^; → ℝ be a gray-scale ICA image with 2D spatial domain Ω, and *g* : Ω → {0, 1} be a binary label mask corresponding to the ground truth: *g*(*p*) = 1 if *p* belongs to the target region *G* and 0 otherwise. Let *s* : Ω → [0, 1] be a pixel-wise softmax output of the base segmentation model. The binary mask *s*^∗^ corresponding to the prediction s is given as:

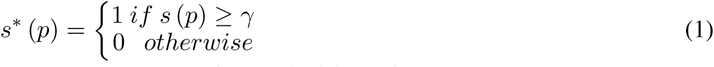

where *γ* is a threshold and the predicted region is defined as *S* = {*p* ∈ Ω | *s*(*p*) ≥ *γ*}.

#### 2.1.1 Dice loss

DSC is a harmonic mean between the ground truth and predicted mask, defined as:

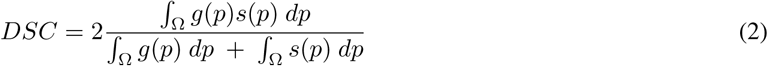

Dice loss *L*_*D*_ is defined as 1-DSC, which is commonly used in semantic segmentation.

#### 2.2.2 Centerline distance loss

Centerline distance loss *L*_*CD*_ evaluates the prediction result using the Euclidean distance between the centerlines of the ground truth and predicted masks, as inspired by Acosta et al. (2017). To obtain the centerline, the soft-skeletonization algorithm of Shit et al. (2021) using iterative min-max pooling on binary masks was applied. Given a binary mask of *y* : Ω → {0, 1}, a recurrent form of soft-skeletonization function can be expressed for *n* ∈ ℕ as:

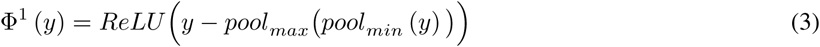

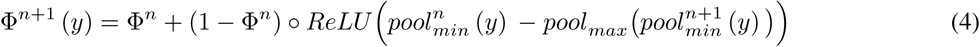

where *pool*_*max*_ and *pool*_*min*_ are shape conservative 3 × 3 max- and min-pooling, respectively, and ○ denotes a Hadamard product. In this study, the maximum iteration of soft-skeletonization was limited to 50 iterations. Accordingly, the binary mask corresponding to the centerline is given as *c* (*y*) = Φ^50^ (*y*) and the centerline is defined as *C* = {*p* ∈ Ω | *c* (*p*) = 1}. A distance map from the centerline C can be defined as:

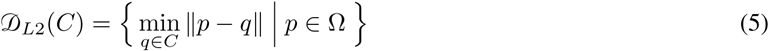

using Euclidean distance transform and modified as:

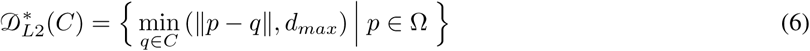

where *d*_*max*_ is a limit of distance to avoid overestimating false positives far from the ground truth. *d*_*max*_ was set as 3. Finally, centerline distance loss is given as:

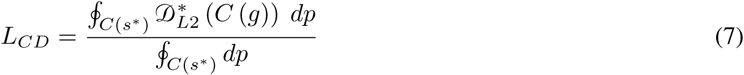

The distance map was precalculated for fast model training.

#### 2.1.3 Edge distance loss

Edge distance loss *L*_*ED*_ evaluates the prediction result by the Euclidean distance between the contours of the ground truth *∂G* and prediction *∂S*, defined as:

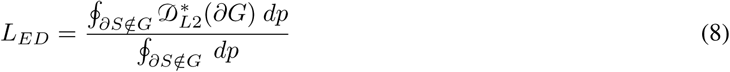

The distance is calculated only outside *∂G* to penalize false positives in the prediction *S*.

#### 2.1.4 Centerline length loss

Centerline length loss *L*_*CL*_ compares the centerline lengths of the ground truth and predicted mask to estimate prediction errors, defined as:

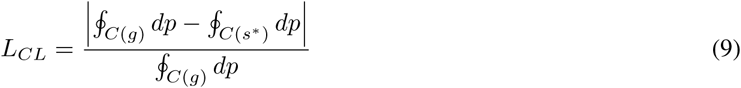

*L*_*CL*_ is an indirect measure to evaluate the concordance of the predicted skeletons for semantic segmentation.

#### 2.1.5 Square dice loss

Square dice loss *L*_*SD*_ has a form of replacing the denominator of *L*_*D*_ to square terms, defined as:

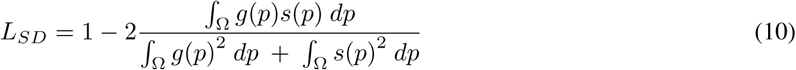

#### 2.1.6 Application of loss functions

*L*_*D*_ and *L*_*SD*_ were assessed independently, whereas *L*_*CD*_, *L*_*ED*_, and *L*_*CL*_ as auxiliary terms were applied while being incorporated with *L*_*D*_. To determine the relative weight of auxiliary losses, a parametric test was performed prior to the cross-validation (Table 1). The relative weights of *L*_*CD*_, *L*_*ED*_, and *L*_*CL*_ with respect to *L*_*D*_ were determined as 0.1, 0.2, and 0.1 that maximized the DSC values, respectively. Consequently, a set of loss functions 𝕃 = {*L*_*D*_, *L*_*D*_ + 0.1*L*_*CD*_, *L*_*D*_ + 0.2*L*_*ED*_, *L*_*D*_ + 0.1*L*_*CL*_, *L*_*SD*_} was determined. Hereafter, the binary mask of the segmentation model with loss function *ℓ* ∈ 𝕃 is indicated as 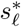.

**Table 1:** Impact of auxiliary loss weight (*α*) on average DSC. Maximum values are presented in bold. *L*_*D*_, Dice loss; *L*_*CD*_, Centerline distance loss; *L*_*ED*_, Edge distance loss; *L*_*CL*_, Centerline length loss.

| $\alpha$ | $L_D + \alpha L_{CD}$ | $L_D + \alpha L_{ED}$ | $L_D + \alpha L_{CL}$ |
| --- | --- | --- | --- |
| 0.1 | **92.35** | 92.12 | **91.83** |
| 0.2 | 92.15 | **92.28** | 91.60 |
| 0.3 | 92.06 | 92.04 | 91.39 |

### 2.2 Selective ensemble methods

#### 2.2.1 Ranking based on morphology

From the source of errors observed in major vessel segmentation (Yang et al., 2019), two hypotheses were established. The first is that the predictive performance is poor if the vascular mask is disconnected. The second is that if a branching pattern is found in the vascular mask, one of the two branches is an obvious error. From the hypotheses, ranking rules for our morphological selective ensemble (MSEN) method were established: 1) rank in order of the centerline length of the major blob; and 2) move the predicted mask with branching patterns to a lower rank. The centerline *C*^∗^ was extracted using the algorithm of Lee et al. (1994) from the python package of scikit-image to avoid errors at the proximal and distal ends of the predicted mask. Branch patterns were detected using the convolution of 3 × 3 kernels with the binary mask *c*^∗^ corresponding to *C*^∗^ (Ardizzone et al., 2008):

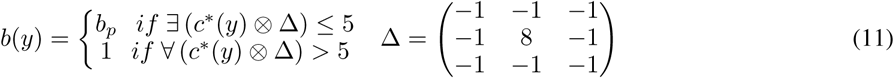

where *b*_*p*_ is a branch penalty, which was set as 2. The rank of base model outcomes is determined in the descending order, using a 5 × 5 exchange matrix *J*:

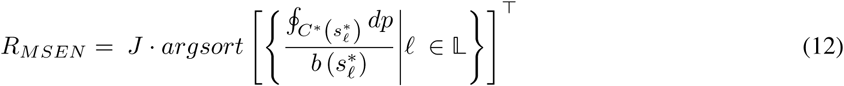

#### 2.2.2 Ranking based on estmiated DSC

The hypothesis for estimated DSC-based selective ensemble (ESEN) was that the pseudo-ground truth from a meta-learner could be used to discriminate a better prediction. By feeding the angiogram and prediction outputs of the validation set, the meta-learner *F*_*meta*_ combining DeepLabV3+ decoder with EfficientNet-B4 encoder was trained (Fig. 1) with Dice loss. The pseudo ground truth *g*^′^ with the same dimension as *g* is defined as:

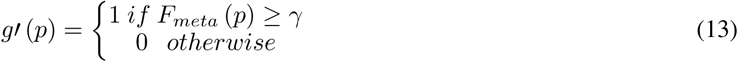

and accordingly, the ranking for ESEN can be expressed as:

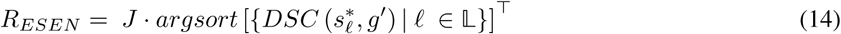

#### 2.2.3 Ensemble output with ranking

During the inference of selective ensemble methods to an image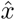, the corresponding final output 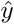 is expressed as:

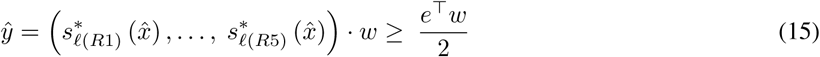

where 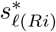 indicates the segmentation output corresponding to the *i*-th ranking, *w* is a column vector for ensemble weights, and *e* is the column vector of ones with the same dimension as *w*. First, *w* = (1, 0, 0, 0, 0) was given to select a single mask with the highest rank, which was indicated as ‘Top-1’. Second, the weight set based on the ranking was optimized by grid search in the integer domain of [0, 10], which maximized the average DSC of the image set and was indicated as ‘WO’ (Fig. 1b).

#### 2.2.4 Hard voting

In an ensemble method, in general, the weight of each model is a fixed constant and does not depend on the prediction outcome of individual images. As a control, hard voting was applied for a pixel-wise ensemble (Fig. 1b). The individual models in hard voting were weighted equally (Hard voting WE) or had weights optimized (Hard voting WO). The weight optimization for hard voting was performed using the grid search as applied for the selective ensemble method.

### 2.3 Quantitative analysis

The segmentation performance was evaluated in two different regions of interest (ROI): the entire image and the local window. The local window was defined as a rectangle of 64 × 64 pixels centered at the minimum lumen diameter of the most narrowed location. If the major vessel had multiple lesions (diameter stenosis >30%), the local window was placed per lesion. To evaluate the performance within the local window, a local DSC was calculated. Considering the imbalance between the vessel area and background, DSC was calculated only for the vessel area.

QCA analysis requires a single mask enclosed by the delineated boundary of the coronary arteries. When the predictive vascular mask consists of multiple blobs, it is not possible to perform QCA analysis without additional image processing to determine whether blobs are placed on the major vessel area and to repair the predicted mask by connecting blobs with an appropriate path. Considering the practical application of deep learning segmentation to automated QCA analysis, further analysis was conducted on the major blob in the same way as the original prediction.

### 2.4 Training setup

Segmentaton models were trained for 400 epochs at maximum with a min-batch size of 12. Input images of 256 × 256 pixels were normalized by using 2D min/max normalization, and the pre-trained weight from ImageNet was adopted for transfer learning. Data augmentation was performed with rotation (−20° to 20°), translation shift (0 to 10% of the image size) and zoom (0 to 10%). The Adam optimizer with *β*1 = 0.9 and *β*2 = 0.999 was applied for training. The learning rate, which was initially set to 10^−3^, halved up to 10^−6^ with the patience of 20 epochs, and the early stop criteria was 80 epochs. The training was conducted using TensorFlow on a workstation with Intel i9-7900X, 128GB RAM, and 4 GeForce GTX 1080Ti.

The prediction network for the pseudo-ground truth of the ESEN method was trained with a mini-batch size of 16. The model input was set as five channels of binary labels from segmentation models concatenated with one channel of the corresponding angiogram. The patience for learning rate decay and the early stop criteria was 40 epochs. The rest of the hyperparameters were identical to those of the segmentation models.

For cross-validation, the internal data was divided into five folds according to the exam date. Each fold contained data from almost the same number of patients, preventing the angiograms of a patient from being included in multiple folds. The fold proportion for training, validation, and evaluation datasets were 3:1:1, and the composition was changed through cyclic permutation in sequence.

### 2.5 Angiography database

#### 2.5.1 Internal dataset

In the present study, 3,309 patients who underwent X-ray coronary angiography in Asan Medical Center from February 2016 to November 2016 were retrospectively enrolled (Fig. 2); the same patients had been enrolled in the previous study for major vessel segmentation (Yang et al., 2019). One angiographic image per major vessel (right coronary artery, RCA; left anterior descending coronary artery, LAD; left circumflex coronary artery, LCX) was selected in the projection angle indicating the lesion morphology (disease vessel with diameter stenosis 30%) or coronary tree structures (normal vessel). Coronary angiograms in which the coronary tree structure was not appropriately recognized, such as chronic total/subtotal occlusion (954 images), poor image quality (incomplete contrast filling, 210; diminutive vessels, 60; blurred images, 88; and severe vessel overlap, 273) and severe overlap with medical device (162 images), were excluded. Finally, a total of 7,426 angiograms from 2,924 patients were included in the internal dataset. With the inclusion of the cases excluded from the previous study (Yang et al., 2019), such as normal and stented vessels, the number of images increased to more than double. The baseline characteristics of the coronary arteries are summarized in Table 2.

**Figure 2:**
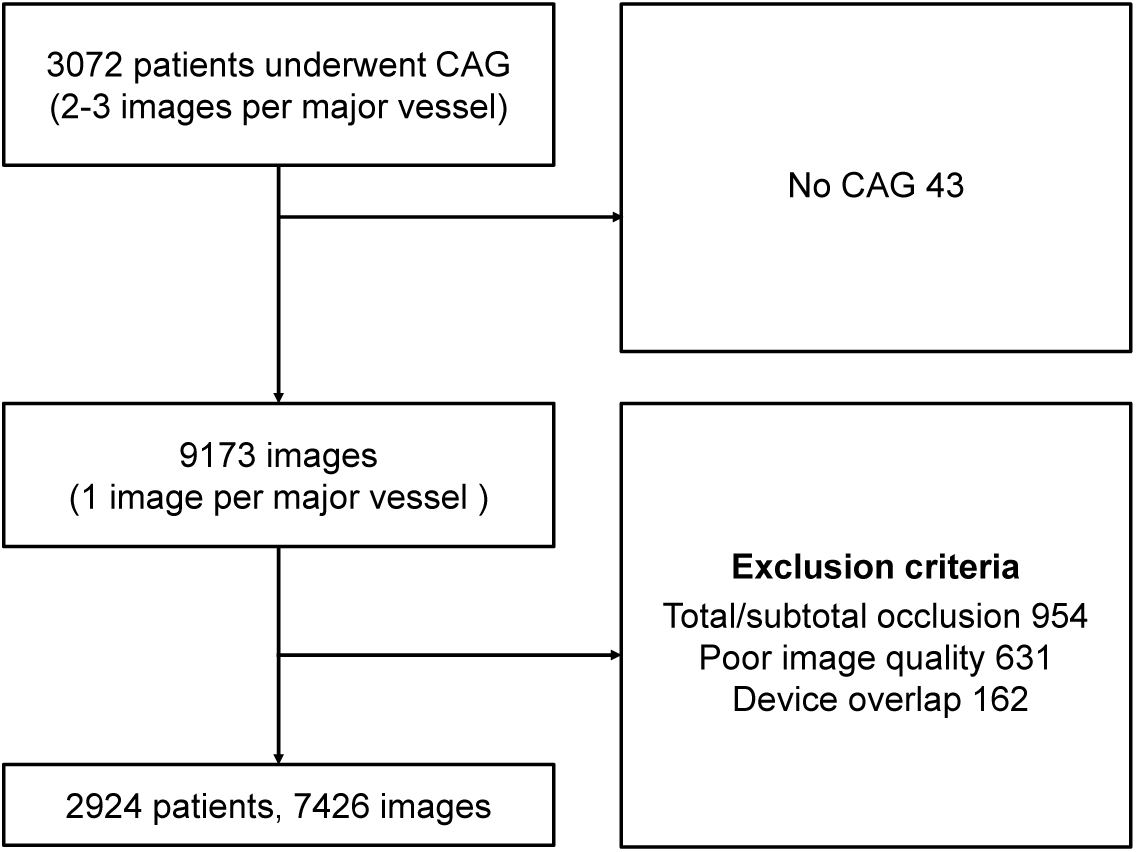
Patient enrollment criteria. After excluding invasive coronary angiography (ICA) in which the coronary tree structure was not appropriately recognized, 7,426 images of 2,924 patients were ultimately included in the present study.

**Table 2:** Angiography statistics. N, number of patients; n, number of images; QCA, quantitative coronary angiography.

| | Internal ($N = 2,924$) | External ($N = 226$) |
| --- | --- | --- |
| Number of vessels, n (%) | 7,426 | 556 |
| Right coronary artery | 2,400 (32.3%) | 175 (31.5%) |
| Left anterior descending artery | 2,546 (34.3%) | 193 (34.7%) |
| Left circumflex artery | 2,480 (33.4%) | 188 (33.8%) |
| Number of diseased vessels, n (%) | 3,896 | 300 |
| Right coronary artery | 1,203 (30.9%) | 92 (30.7%) |
| Left anterior descending artery | 1,678 (43.1%) | 127 (42.3%) |
| Left circumflex artery | 1,015 (26.0%) | 81 (27.0%) |
| % diameter stenosis | $46.2 \pm 15.0$ | $47.7 \pm 13.9$ |
| Lesion length (mm) | $18.2 \pm 10.7$ | $17.9 \pm 11.0$ |

#### 2.5.2 External dataset

The deep learning models trained with the internal dataset were further assessed using the external dataset. The external dataset consisted of 556 images of 226 patients who visited Chungnam National University Hospital from February 2016 to November 2016. The protocol for the enrollment and exclusion of angiographic images was the same as that for the internal dataset.

#### 2.5.3 Labeling process

For the labeling process, angiograms in DICOM file format were anonymized by removing patient-identifiable information with the dedicated tool provided by the Big Data Research Center of Asan Medical Center. Using CAAS Workstation 7.5 (Pie Medical Imaging BV, Maastricht, Netherlands), three expert radiographers with more than 10 years of experience each annotated the major vessel areas. Each label was generated on the image frame at end-diastole by a semi-automatic edge-detection tool. The vessel area was then manually corrected. The target area of major vessel segmentation was set as the region from the ostium to the far distal location. Pixel information of the major vessel area was extracted using a customized Python script.

This study complies with the Declaration of Helsinki. Research approval was granted by the Institutional Review Board of Asan Medical Center and Chungnam National University Hospital. The requirement for informed consent from the patients was waived.

### 2.6 Statistical analysis

Continuous variables are presented as mean ś standard deviation (SD), and categorical variables are presented as numbers and percentages. Linear regression analysis was performed to estimate the relationship of the maximum difference in DSCs with the average DSC from the individual models. The Wilcoxon rank sum test (Mann-Whitney U test) was conducted to compare the segmentation performance between pairwise ranking in MSEN and ESEN. p values <0.05 were considered statistically significant, and the normality of data was tested with the Shapiro-Wilk W-test (p>0.05). All of the statistical analyses were performed using SPSS, Version 17.0 for Windows (IBM Corp., Armonk, NY, USA).

## 3 Results

### 3.1 Segmentation performance of individual models

The segmentation models exhibited a minor difference <0.11% except for the combination of Dice and Centerline length losses (*L*_*D*_ + 0.1*L*_*CL*_) with a relatively low DSC (Table 3). However, the segmentation outputs demonstrated different morphological responses to the loss functions (Fig. 3). The segmentation model, which provided an accurate prediction of major vessel areas, differed from image to image, and the DSC that could be achieved varied depending on the model applied. The maximum differences in DSCs between individual models were greater for images with a lower mean DSC (*y* = −0.593 *x* + 0.591, *R*^2^ = 0.340 in Fig. 4a). The probabilities of individual models that performed best for images with a mean DSC <90% showed little deviation (15.74% –21.83% in Fig. 4b).

**Table 3:** Average DSCs (%) of individual and ensemble models from internal dataset. Highest scores along the column are highlighted in bold. *L*_*D*_, Dice loss; *L*_*CD*_, Centerline distance loss; *L*_*ED*_, Edge distance loss; *L*_*CL*_, Centerline length loss; *L*_*SD*_, Square dice loss; WE, weighted equally; WO weight optimized.

| Method | DSC |  | Local DSC |  |
| --- | --- | --- | --- | --- |
|  | Original | Major blob | Original | Major blob |
| **Individual model** |  |  |  |  |
| $L_D$ | 92.49 ± 8.34 | 92.07 ± 9.71 | 93.35 ± 12.85 | 92.56 ± 15.25 |
| $L_D + 0.1L_{CD}$ | 92.57 ± 8.17 | 92.14 ± 9.38 | 93.40 ± 12.73 | 92.57 ± 15.03 |
| $L_D + 0.2L_{ED}$ | 92.46 ± 8.39 | 91.90 ± 9.89 | 93.14 ± 13.38 | 92.28 ± 15.62 |
| $L_D + 0.1L_{CL}$ | 92.12 ± 8.05 | 91.59 ± 9.49 | 92.86 ± 12.80 | 92.01 ± 15.11 |
| $L_{SD}$ | 92.54 ± 7.97 | 92.10 ± 9.41 | 93.45 ± 12.42 | 92.64 ± 14.92 |
| **Simple ensemble model** |  |  |  |  |
| Hard voting WE | 93.06 ± 7.66 | 92.60 ± 9.01 | 93.84 ± 12.07 | 93.08 ± 14.36 |
| Hard voting WO | 93.10 ± 7.46 | 92.72 ± 8.75 | 93.98 ± 11.63 | 93.29 ± 13.85 |
| **Simple ensemble model** |  |  |  |  |
| MSEN Top-1 (Proposed) | 92.59 ± 8.35 | 92.48 ± 8.98 | 93.76 ± 12.11 | **93.55 ± 12.95** |
| MSEN WO (Proposed) | 93.10 ± 7.61 | 92.78 ± 8.74 | 94.04 ± 11.59 | 93.52 ± 13.33 |
| ESEN Top-1 (Proposed) | 92.88 ± 7.80 | 92.56 ± 8.95 | 93.81 ± 11.96 | 93.18 ± 13.91 |
| ESEN WO (Proposed) | **93.11 ± 7.61** | **92.80 ± 8.69** | **94.04 ± 11.54** | 93.49 ± 13.37 |
| Highest DSC among 5 individual models | 94.30 ± 5.85 | 94.18 ± 6.53 | 95.39 ± 8.78 | 95.10 ± 10.13 |

**Figure 3:**
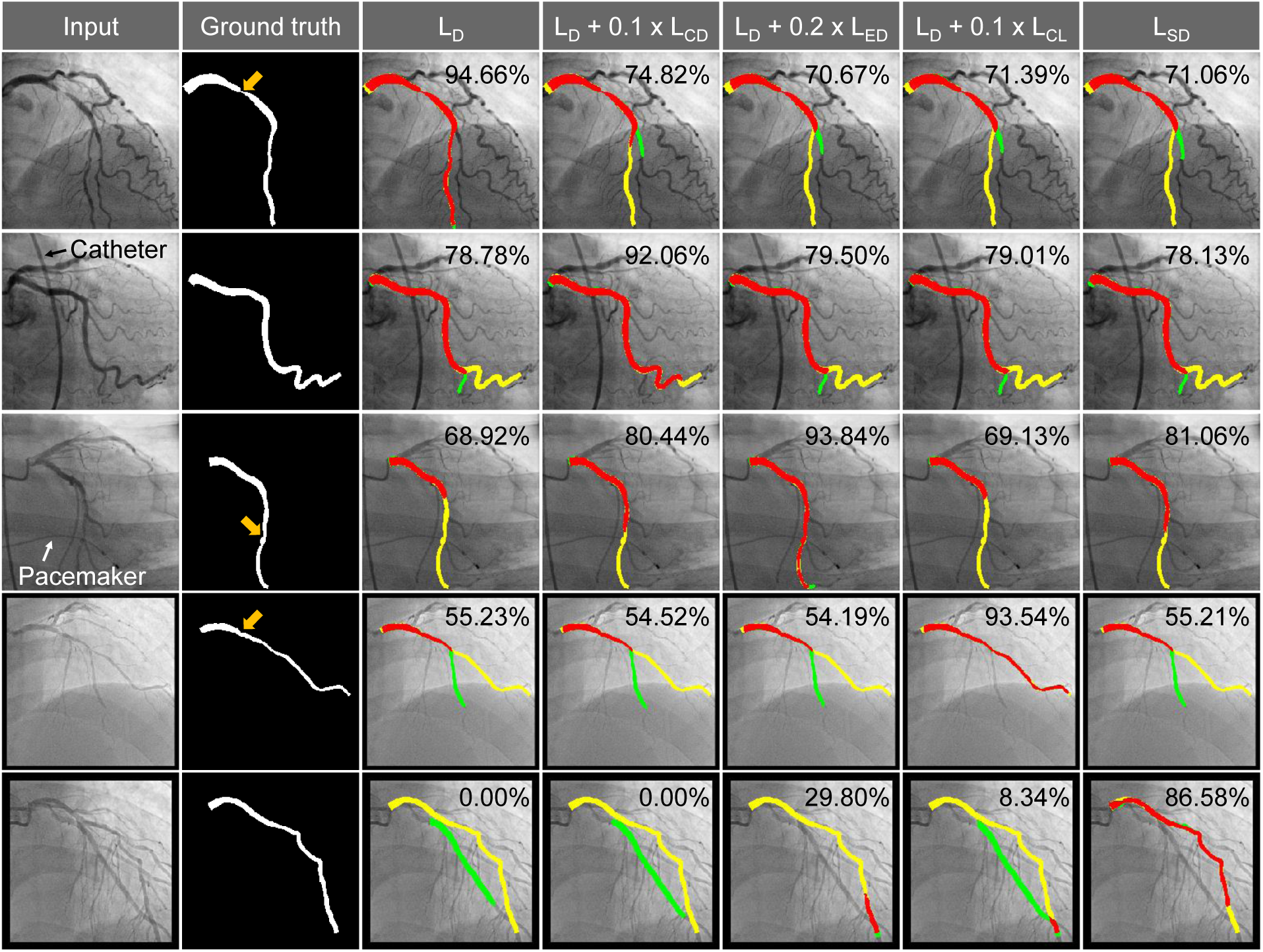
Representative examples of major vessel segmentation from 5 individual models. In the third to seventh columns, the predicted major vessel areas compared to the ground truth (second column) are indicated in red (true positive), yellow (false negative), and green (false positive). The DSC for each model is shown in the upper right corner. Orange arrows in the second column indicate coronary lesions. *L*_*D*_, Dice loss; *L*_*CD*_, Centerline distance loss; *L*_*ED*_, Edge distance loss; *L*_*CL*_, Centerline length loss; *L*_*SD*_, Square dice loss

**Figure 4:**
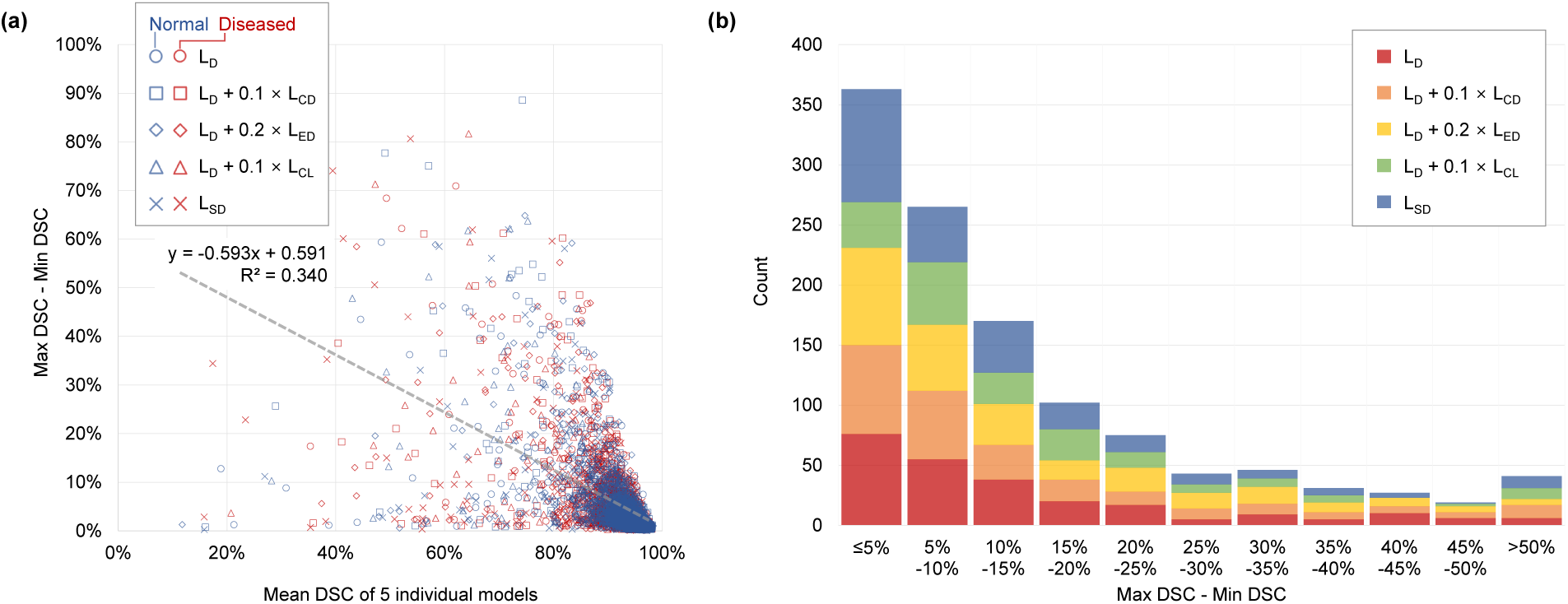
Performance analysis in relation to DSC variances. (a) Scatter plot for the difference between the maximum and minimum DSCs versus Mean DSC. Symbol indicates the segmentation model with the highest DSC. (b) The difference between the maximum and minimum DSCs is plotted as a histogram for images with mean DSC of less than 90% for five individual models. The color indicates the segmentation model with the highest DSC. DSC, dice similarity coefficient; *L*_*D*_, Dice loss; *L*_*CD*_, Centerline distance loss; *L*_*ED*_, Edge distance loss; *L*_*CL*_, Centerline length loss; *L*_*SD*_, Square dice loss.

### 3.2 Ranking for selective ensemble methods and weight optimization

The ranking of the MSEN and ESEN methods successfully distinguished better outcomes among the outputs from individual models (Fig. 5). In the MSEN method, images in the top three ranks had statistically higher DSCs than did lower-rank images. In the ESEN method, the pseudo-ground truth from the meta-learner showed a slight improvement in average DSC (92.58% ± 7.39%) but successfully discriminated better predictions. That is, the higher the ranking, the better the segmentation performance, except for the 1st versus 2nd ranks. Therefore, in MSEN WO and ESENWO models, images with higher ranking had greater weights of (4, 3, 3, 2, 2) and (2, 2, 2, 1, 1), respectively. Meanwhile, the optimized weight in the Hard voting WO model was (3, 2, 2, 3, 4) for the loss function *ℓ* ∈ 𝕃.

**Figure 5:**
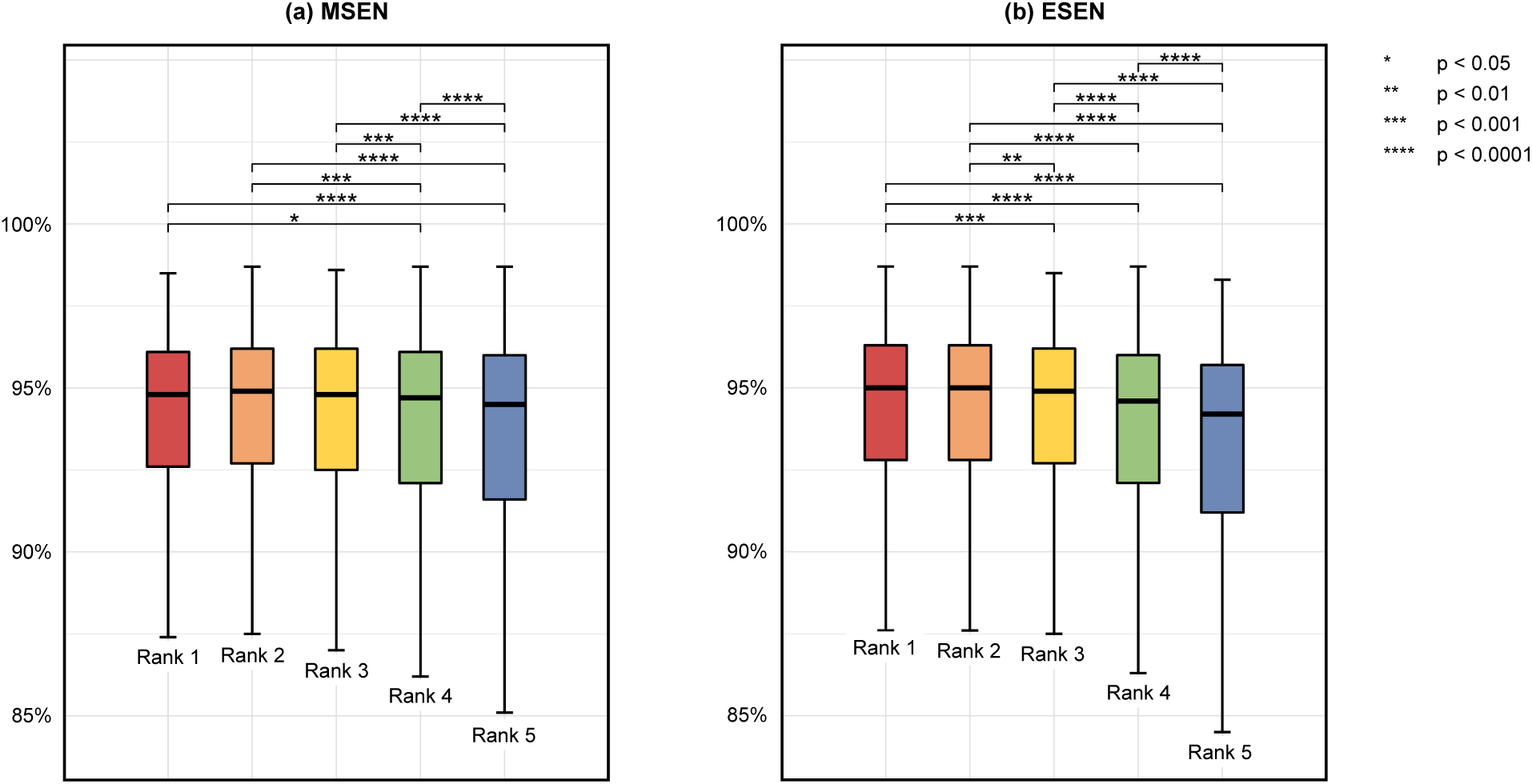
Comparison of DSCs according to the ranking determined by (a) MSEN and (b) ESEN methods. DSC, dice similarity coefficient; MSEN, morphological selective ensemble; ESEN, estimated dice similarity coefficient-based selective ensemble.

### 3.3 Segmentation performance of selective ensemble models

All of the analyzed selective ensemble models improved the average DSCs compared to the individual models (Table 3). The ESEN WO model produced the best DSC of 93.11% ± 7.61%, followed by the MSEN WO and Hard voting WO models. Near the most narrowed region of the major vessel, the ESEN WO model also had the best performance (local DSC = 94.04% ± 11.59%). The improvement of DSC achieved by selective ensemble models was greater in the local window (0.31% –1.18%) than in the entire image (0.02% –0.99%).

Even when disconnection occurred in the majority of individual models, selective ensemble models provided an accurate prediction for major vessel areas (Fig. 6a). Although MSEN Top-1 successfully selected an outcome with better connectivity near the lesion (Fig. 6b), the ranking of the MSEN method was susceptible to false positives in the longitudinal direction of major vessels (Fig. 6c). Compared to the MSEN WO models, the ESEN WO model, which gave greater weights to images with relatively higher ranking, produced better results by combining poor prediction regions (Fig. 6d).

**Figure 6:**
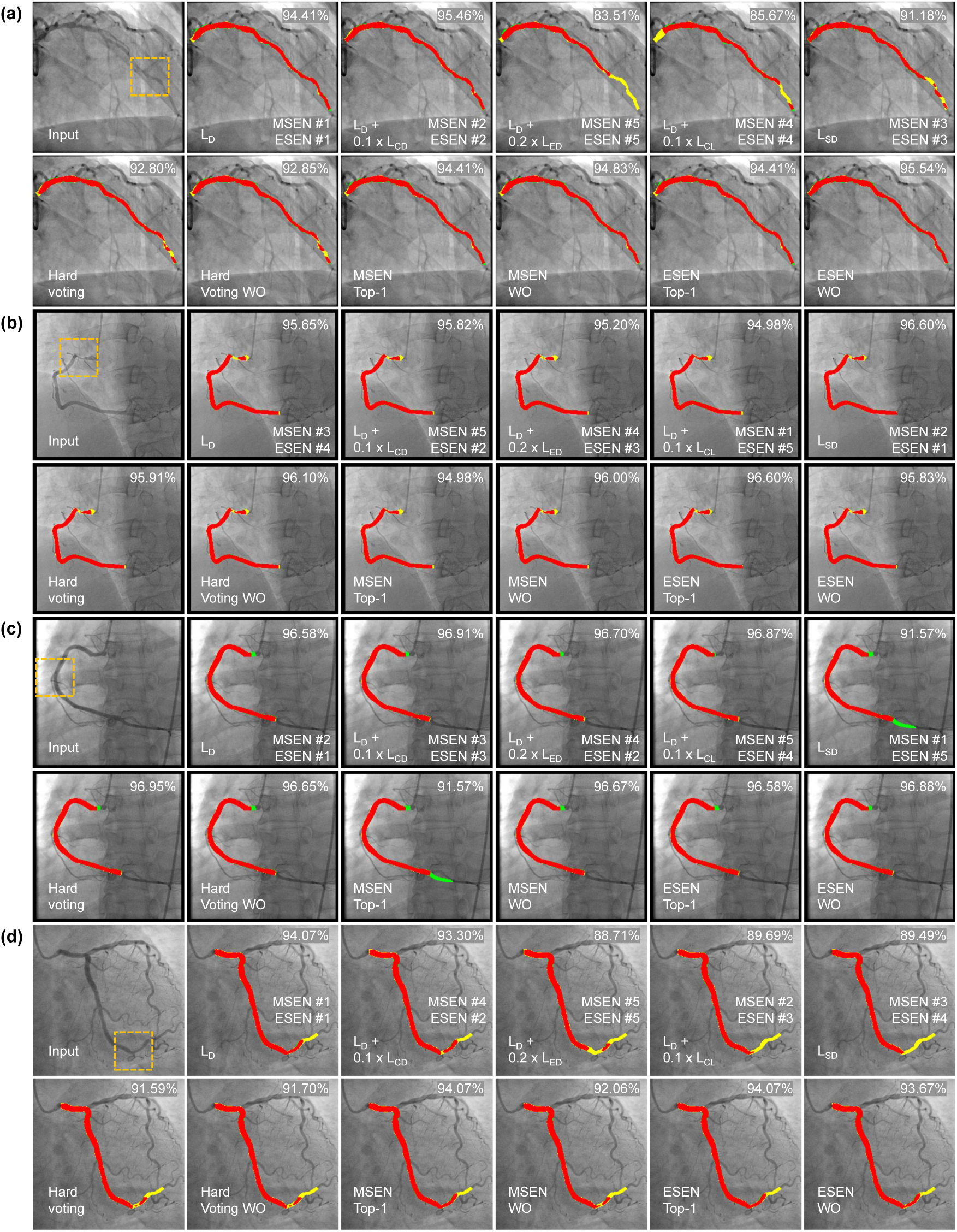
Representative examples for comparison of individual and ensemble models. The predicted major vessel areas compared to the ground truth are indicated in red (true positive), yellow (false negative), and green (false positive). The DSC for each model is shown in the upper right corner. The ranking in MSEN and ESEN methods are presented in the lower right corner. The orange box in the input image indicates the local window near the most narrowed region. DSC, dice similarity coefficient; ESEN, estimated dice similarity coefficient-based selective ensemble; MSEN, morphological selective ensemble; *L*_*D*_, Dice loss; *L*_*CD*_, Centerline distance loss; *L*_*ED*_, Edge distance loss; *L*_*CL*_, Centerline length loss; *L*_*SD*_, Square dice loss; WE, weighted equally; WO weight optimized.

### 3.4 Major blob analysis

Since the disconnected blobs were likely to be distributed on the major vessel area, their removal resulted in the DSC decrease (Table 3). For individual models, the DSC reduction was more conspicuous in the local window (0.79% –0.87%) compared to the entire image (0.42% –0.56%). Selective ensemble models were less affected by disconnected blob exclusion than were the individual and hard voting models. The MSEN Top-1 model minimized the difference in DSCs between the original prediction and major blob (−0.11% for the entire image; -0.21% for the local window), indicating fewer occurrences of mask disconnection. For the major blob in the local window, MSEN models outperformed other segmentation models.

### 3.5 Cumulative histogram

With a selective ensemble model, the proportion of images exhibiting DSC >90% was more than 86.43% in the original prediction, whereas the highest among the individual models was 85.01% with *L*_*D*_ + 0.1*L*_*CD*_ (Table 4 and Fig. 7). In the local window, all of the selective ensemble models were more likely to have DSC >90% than the hard voting models, and their difference was maximum for the major blob in the local window (90.39% for MSEN Top-1 vs. 88.84% for Hard voting WE).

**Table 4:** Average DSCs (%) of individual and ensemble models from internal dataset. Highest scores along the column are highlighted in bold. *L*_*D*_, Dice loss; *L*_*CD*_, Centerline distance loss; *L*_*ED*_, Edge distance loss; *L*_*CL*_, Centerline length loss; *L*_*SD*_, Square dice loss; WE, weighted equally; WO weight optimized.

| Method | DSC >90% |  | Local DSC >90% |  |
| --- | --- | --- | --- | --- |
|  | Original | Major blob | Original | Major blob |
| **Individual model** |  |  |  |  |
| $L_D$ | 84.89 | 84.28 | 88.11 | 87.68 |
| $L_D + 0.1L_{CD}$ | 85.01 | 84.10 | 88.55 | 87.89 |
| $L_D + 0.2L_{ED}$ | 84.62 | 83.54 | 88.45 | 87.53 |
| $L_D + 0.1L_{CL}$ | 83.96 | 83.22 | 87.95 | 87.26 |
| $L_{SD}$ | 84.89 | 84.34 | 88.67 | 88.23 |
| **Simple ensemble model** |  |  |  |  |
| Hard voting WE | 87.25 | 86.22 | 89.66 | 88.84 |
| Hard voting WO | 87.34 | 86.71 | 89.79 | 89.20 |
| **Simple ensemble model** |  |  |  |  |
| MSEN Top-1 (Proposed) | 86.43 | 86.61 | 90.43 | 90.39 |
| MSEN WO (Proposed) | 87.72 | 87.25 | 90.20 | 89.85 |
| ESEN Top-1 (Proposed) | 86.92 | 86.40 | 89.97 | 89.54 |
| ESEN WO (Proposed) | 87.61 | 87.29 | 90.04 | 89.63 |

**Figure 7:**
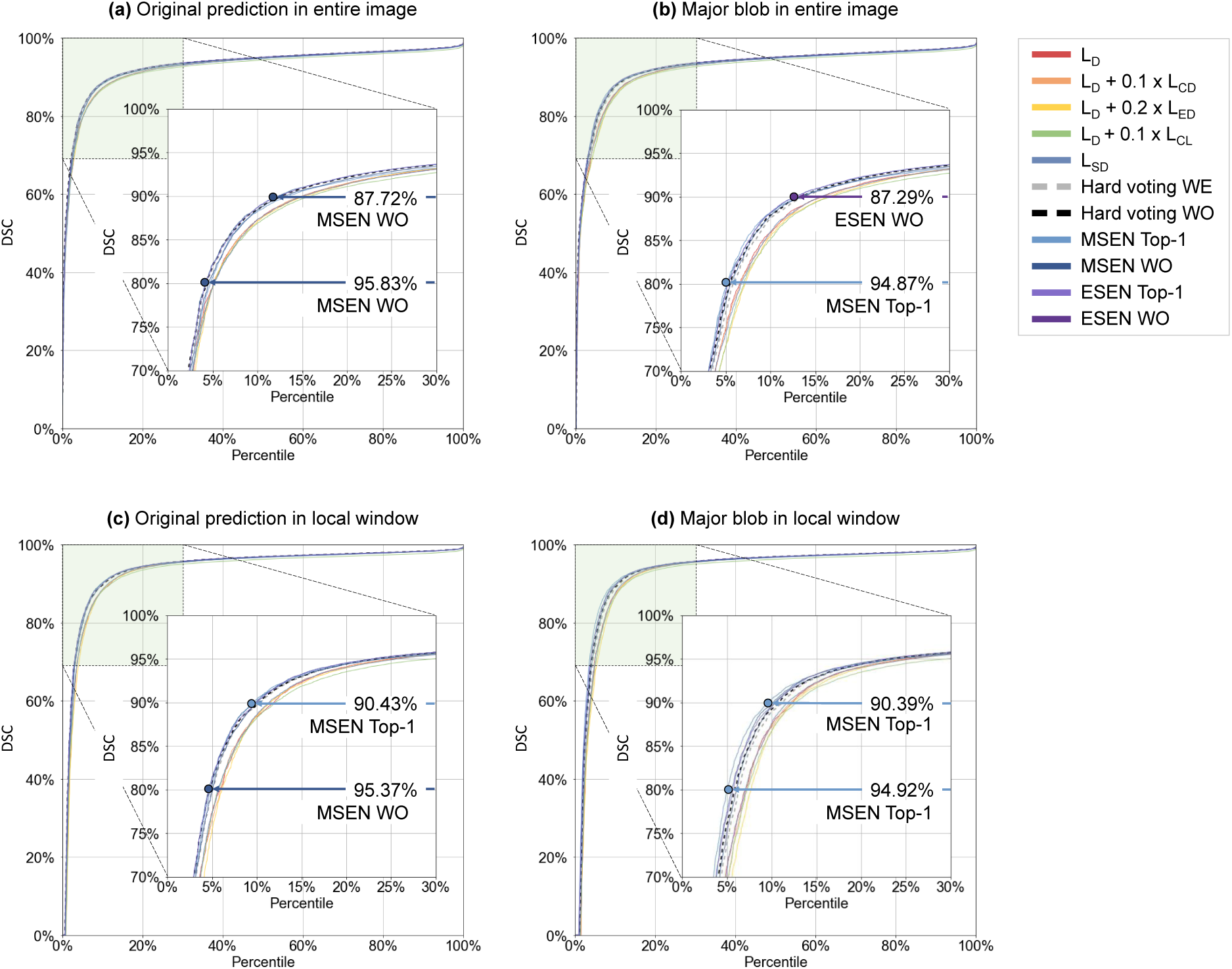
Cumulative histograms of DSCs. The maximum proportions of images with DSCs > 80% and % are indicated with circles and arrows. DSC, dice similarity coefficient; ESEN, estimated dice similarity coefficient-based selective ensemble; MSEN, morphological selective ensemble; *L*_*D*_, Dice loss; *L*_*CD*_, Centerline distance loss; *L*_*ED*_, Edge distance loss; *L*_*CL*_, Centerline length loss; *L*_*SD*_, Square dice loss; WE, weighted equally; WO weight optimized.

### 3.6 Error analysis

Selective ensemble models reduced the segmentation errors of mask disconnection compared to the individual models (Fig. 8). With the MSEN Top-1 model, the mask disconnection was reduced to less than three-fifths of the predictive model with *L*_*D*_, and the discontinuity at the most narrowed region occurred in <1% of the tested images (13/1457). Among the individual models, the loss function of *L*_*D*_ + 0.1*L*_*CL*_ used to evaluate the centerline length showed better connectivity. Misidentification error slightly improved in the ensemble models.

**Figure 8:**
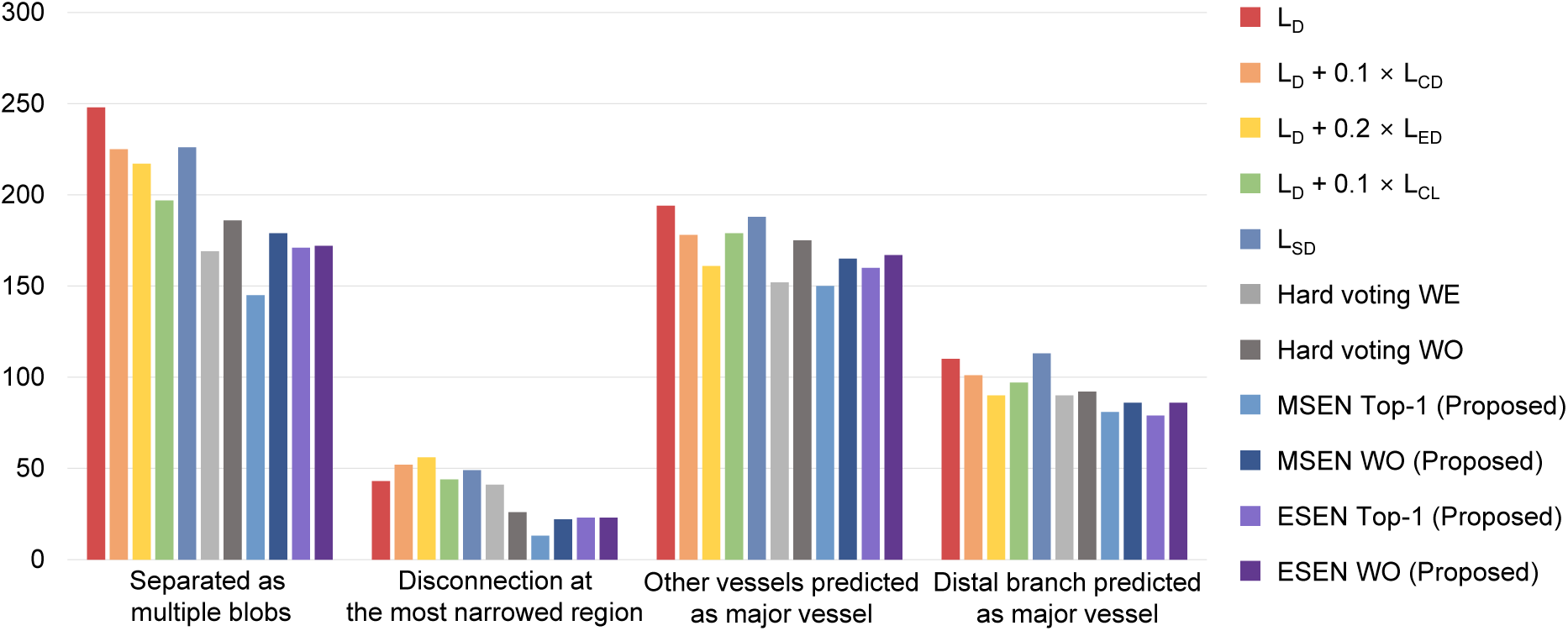
Error analysis of segmentation models for each error type. Two types of segmentation errors, disconnection and misidentification, were classified in a fold of the internal dataset (1,457 images). MSEN, morphological selective ensemble; *L*_*D*_, Dice loss; *L*_*CD*_, Centerline distance loss; *L*_*ED*_, Edge distance loss; *L*_*CL*_, Centerline length loss; *L*_*SD*_, Square dice loss; WE, weighted equally; WO weight optimized.

### 3.7 External validation

In the external dataset, the combination of Dice and Centerline length losses (*L*_*D*_ + 0.1*L*_*CL*_) performed better than other individual models did, except for the original prediction of the entire image in which *L*_*SD*_ had the highest DSC (Table 5). The MSEN and ESEN models, as in the internal dataset, enhanced predictive capabilities in all of the ROIs. The benefits of selective ensemble models were apparent in the local window. In the major blob analysis of the local window, MSEN Top-1 models improved the DSC by more than 1.23% (vs. hard voting models) and by more than 2.08% (vs. individual models).

**Table 5:**
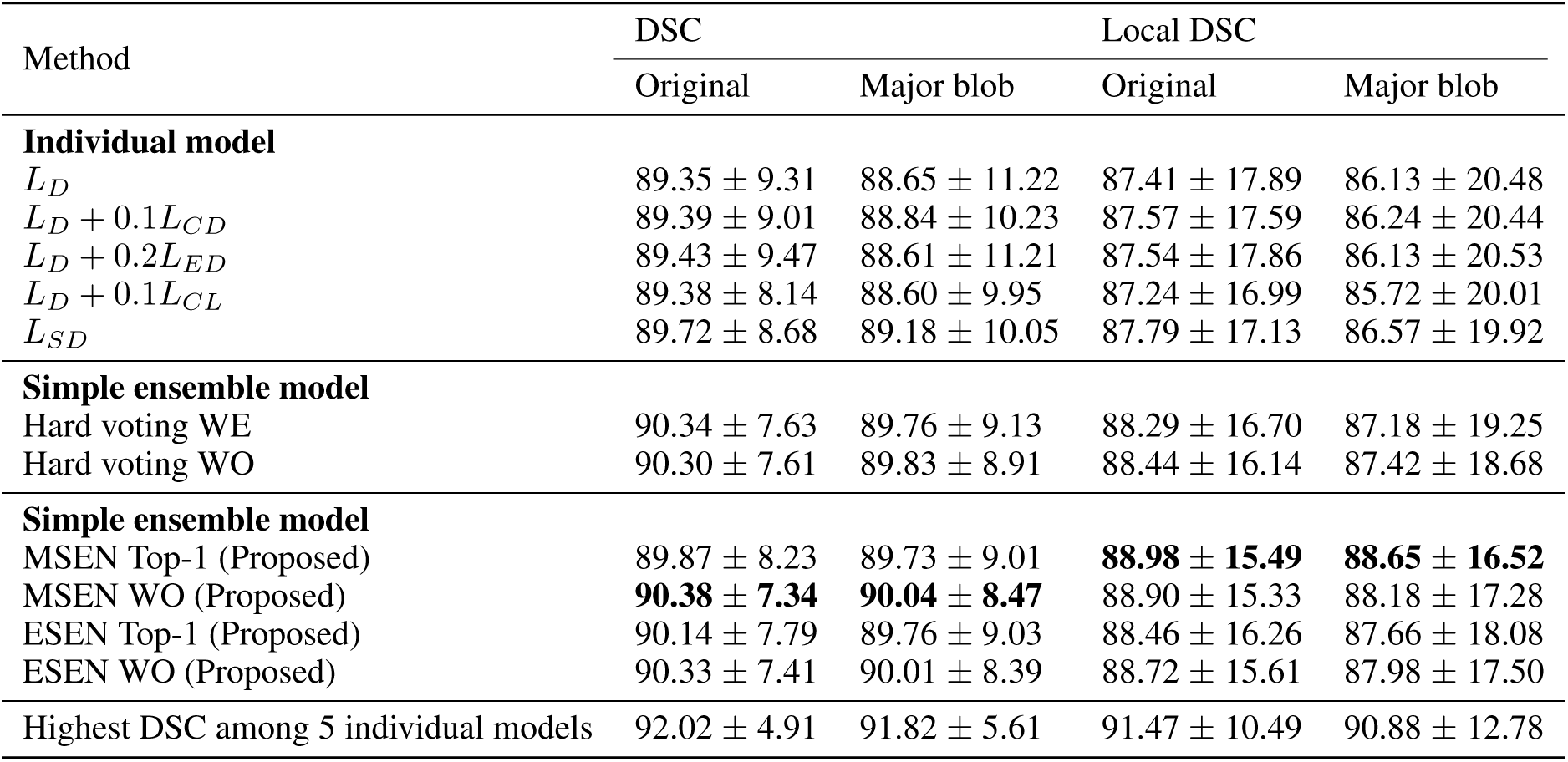
Average DSCs (%) of individual and ensemble models from external dataset. Highest scores along the column are highlighted in bold. *L*_*D*_, Dice loss; *L*_*CD*_, Centerline distance loss; *L*_*ED*_, Edge distance loss; *L*_*CL*_, Centerline length loss; *L*_*SD*_, Square dice loss; WE, weighted equally; WO weight optimized.

## 4 Discussion and conclusion

In the present study, we proposed two selective ensemble methods in which the weights on prediction outcomes were imposed differently to individual images rather than fixed according to the average performance of individual models. The selective ensemble methods improved the segmentation performance, with DSCs of up to 93.11% in the entire image, and enabled a more accurate delineation of the coronary lesion. Here, a DSC >90% was achieved by 87.72% of the image dataset, and the probability of mask disconnection at the most narrowed region could be minimized to <1%. For the external dataset with different image characteristics, although the superiority between the average performance of individual models was changed, the MSEN and ESEN methods exhibited less performance degradation.

Our selective ensemble models demonstrated an accurate prediction of the major vessel in coronary angiography. Compared to previous studies for major vessel segmentation (Jun et al., 2020; Yang et al., 2019), the average DSC was increased by more than 1.41%. The proportion of images with DSC <90% was also reduced to about two-thirds (Yang et al., 2019). The previous studies used a single segmentation model trained using a smaller dataset (44.5% –63.3% of our database). Yang et al. (2019) achieved a DSC of 91.7% for only diseased coronary vessels. Unlike previous studies (Jun et al., 2020; Yang et al., 2019), the inclusion of the left main coronary artery into the label, where vessel overlap occurs frequently, allowed the detection of the ostium lesions of LAD and LCX. DSCs >90% for the external dataset indicated greater applicability for general use. Recently updated deep learning studies for the segmentation of the entire coronary tree exhibited DSCs of 86.4% –88% using a dataset of <1,000 patients (Iyer et al., 2021; Zhao et al., 2018). Despite the detailed capture of vascular structures, entire vessel segmentation could suffer from small branches (Wang et al., 2020). Because the segmentation criteria and the database may vary between studies (Iyer et al., 2021; Wang et al., 2020; Zhao et al., 2018), the interpretation of evaluation metrics should be made with careful consideration.

The MSEN method was inspired by clinical intuition with observation. Human anatomy in medical imaging has a set form, and medical experts could recognize deficiencies in the prediction by observing the segmentation outputs. In major vessel segmentation, the simple rule of the predicted mask should be not disconnected and branched worked well as an evaluation criterion for ranking. Instead of this empirical approach, anomaly detection (Liu et al., 2019; Sandfort et al., 2021; Schlegl et al., 2017; Xia et al., 2020) would be helpful in discriminating plausible candidates for the ensemble. In the post-processing stage, the morphology-based ranking could be corrected through quantitative analysis or allometric laws governing coronary anatomy (Huo and Kassab, 2009). For other organs in medical imaging, registration with the reference atlas that has been used for the segmentation of medical imaging (Išgum et al., 2009; Lorenzo-Valdés et al., 2002) could be employed for evaluation.

The ESEN method involved the meta-learner that was trained using prediction results from individual modes as an input. Unlike the previous studies in which the meta-learner’s outcome was determined as the final output (Zheng et al., 2019), the ESEN method used it as a pseudo-ground truth for quality control. In our application of the ESEN method, the performance of the individual prediction selected based on the pseudo-ground truth was better than the pseudo-ground truth. Although the pseudo-ground truth predicted a mask close to the ground truth, it also partly included minor predictions that were likely false positives. Therefore, it was not practical to select the pseudo-ground truth as the final output. Although both methods premised that the average prediction performance was sufficiently high, the ESEN method differed in that the ranking criterion using the pseudo-ground truth was affected by individual predictions. This method could be more susceptible to different image characteristics at the deployment stage, as shown in the local window analysis of external validation.

Recent developments in automated segmentation methods for ICA images could lead to changes in the clinical activity of coronary intervention. First, real-time analysis in the catheterization room could help guide and optimize the stent selection and implantation process beyond error-prone visual assessment (Shah et al., 2017). Automatic lesion detection integrated with segment identification (Zhai et al., 2019) would facilitate the diagnosis of multivessel disease with the calculation of the SYNTAX score (Cavalcante et al., 2017). Machine learning applications using lesion morphology for the prediction of fractional flow reserve could be accelerated (Cho et al., 2019). In applying computational fluid dynamics to 3D QCA, the bottleneck of geometric reconstruction could be tackled to estimate functional diagnostic values (Asano et al., 2019; Ono et al., 2021) and plaque vulnerability (Kweon et al., 2018; Wu et al., 2020). The time required to apply selective ensemble methods (0.36 s in Table 6) would not delay the clinical practice considering the overall duration of the coronary intervention (approximately 30 mins), facilitating the novel techniques using coronary angiography.

**Table 6:**
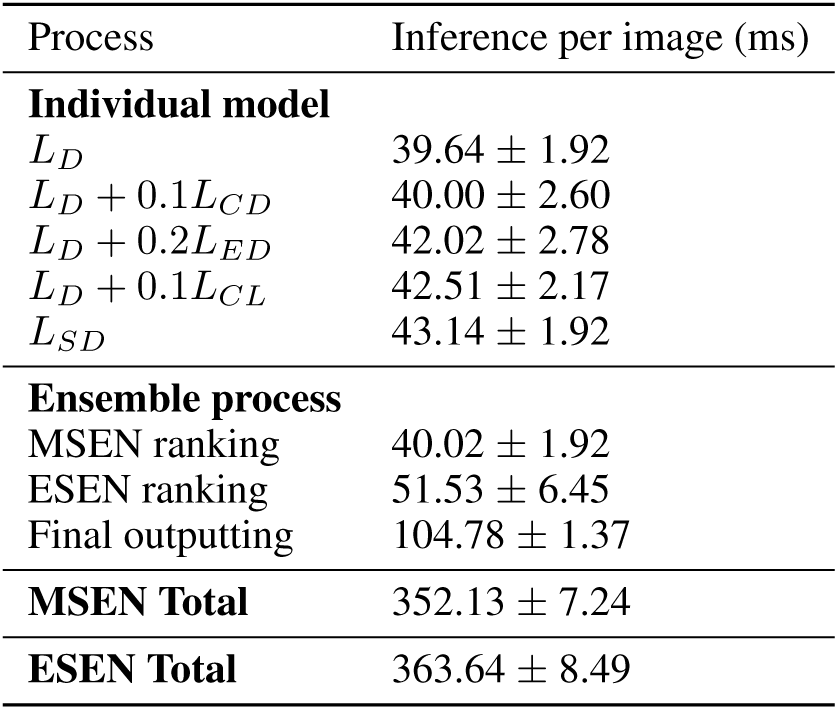
Computational cost to apply ensemble models. MSEN, Morphological selective ensemble; ESEN, Estimated dice similarity coefficient-based selective ensemble; *L*_*D*_, Dice loss; *L*_*CD*_, Centerline distance loss; *L*_*ED*_, Edge distance loss; *L*_*CL*_, Centerline length loss; *L*_*SD*_, Square dice loss.

For the development of fully automated QCA, our approach had some aspects that need to be improved on. Although the proportion of major vessels in coronary lesions was dominant (Halon et al., 1983), side branch analysis is necessary to understand the coronary physiology and evaluate the bifurcation lesion. Analysis across angiographic sequences is necessary for adequate frame selection and more comprehensive interpretation. Since the performance of selective ensemble methods was bound to the best scores of individual models (Tables 3 and 5), an optimal combination of improved individual models is a prerequisite for a better performance of the selective ensemble models. In addition, to assess the performance robustness of our segmentation models, further validation studies should be conducted for the diversity of angiography characteristics. In summary, we proposed selective ensemble methods that ranked the prediction results obtained from multiple segmentation models and imposed greater weights to images with higher rankings. The evaluation criteria for ranking, morphology, and estimated DSC of predicted masks were applied. In the MSEN method, the ranking criteria were empirically determined to avoid the frequent types of segmentation errors. In the ESEN method, a deep neural network was introduced as a meta-learner for the generation of pseudo-ground truth. The selective ensemble methods improved the segmentation performance in the entire image area and provided better delineation around the lesion. Therefore, by enabling more automated QCAs to minimize the clinicians intervention, our selective ensemble methods may allow the real-time application of QCA-based diagnostic methods in routine clinical settings.

## Data Availability

The datasets generated and/or analyzed during the current study are not publicly available because the permission of sharing patient data was not granted by the Institutional Review Board but are available from the corresponding author on reasonable request.

## Acknowledgements

This material was supported by the Institute of Information & Communications Technology Planning & Evaluation (IITP) grant funded by the Korea government (MSIT) (No. 2020-0-00159), and by the Korea Medical Device Development Fund grant funded by the Korea government (the Ministry of Science & ICT, the Ministry of Trade, Industry & Energy, the Ministry of Health & Welfare, the Ministry of Food and Drug Safety) (Project Number: KMDF_PR_20200901_0013).

## Author Contributions

**Jeeone Park**: Methodology, Software, Validation, Writing - Original Draft **Jihoon Kweon**: Conceptualization, Writing - Original Draft, Supervision, Funding acquisition **Hyehyeon Bark**: Methodology, Software **Young In Kim, Inwook Back, Jihye Chae**: Formal analysis **Jae-Hyung Roh, Do-Yoon Kang, Pil Hyung Lee, Jung-Min Ahn, Soo-Jin Kang, Duk-Woo Park, Seung-Whan Lee, Cheol Whan Lee, Seong-Wook Park, Seung-Jung Park**: Resources, Writing - Review & Editing **Young-Hak Kim**: Writing - Review & Editing, Supervision

## References

O. Acosta, E. Mylona, M. Le Dain, C. Voisin, T. Lizee, B. Rigaud, C. Lafond, K. Gnep, and R. de Crevoisier. Multi-atlas-based segmentation of prostatic urethra from planning CT imaging to quantify dose distribution in prostate cancer radiotherapy. Radiotherapy and Oncology, 125(3):492–499, 2017. ISSN 18790887.

E. Ardizzone, R. Pirrone, O. Gambino, and S. Radosta. Blood vessels and feature points detection on retinal images. In Proceedings of the 30th Annual International Conference of the IEEE Engineering in Medicine and Biology Society, EMBS’08 - “Personalized Healthcare through Technology”, pages 2246–2249, 2008. ISBN 9781424418152.

T. Asano, Y. Katagiri, C. C. Chang, N. Kogame, P. Chichareon, K. Takahashi, R. Modolo, E. Tenekecioglu, C. Collet, H. Jonker, C. Appleby, A. Zaman, N. van Mieghem, N. Uren, J. Zueco, J. J. Piek, J. H. Reiber, V. Farooq, J. Escaned, A. P. Banning, P. W. Serruys, and Y. Onuma. Angiography-Derived Fractional Flow Reserve in the SYNTAX II Trial: Feasibility, Diagnostic Performance of Quantitative Flow Ratio, and Clinical Prognostic Value of Functional SYNTAX Score Derived From Quantitative Flow Ratio in Patients With 3-Vessel Dis. JACC: Cardiovascular Interventions, 12(3):259–270, 2019. ISSN 18767605.

M. Baldeon Calisto and S. K. Lai-Yuen. AdaEn-Net: An ensemble of adaptive 2D3D Fully Convolutional Networks for medical image segmentation. Neural Networks, 126:76–94, 2020. ISSN 18792782.

C. Blondel, G. Malandain, R. Vaillant, and N. Ayache. Reconstruction of coronary arteries from a single rotational x-ray projection sequence. IEEE Transactions on Medical Imaging, 25(5):653–663, 2006.

G. Brown, J. Wyatt, R. Harris, and X. Yao. Diversity creation methods: A survey and categorisation. Information Fusion, 6(1):5–20, 2005. ISSN 15662535.

N. Byrne, J. R. Clough, G. Montana, and A. P. King. A Persistent Homology-Based Topological Loss Function for Multi-class CNN Segmentation of Cardiac MRI. In Springer Nature Switzerland AG, volume 12592 LNCS, pages 3–13, Cham, 2021. Springer International Publishing. ISBN 9783030681067.

F. Caliva, C. Iriondo, A. M. Martinez, S. Majumdar, and V. Pedoia. Distance Map Loss Penalty Term for Semantic Segmentation. arXiv preprint, page 1908.03679, 2019. ISSN 2331-8422.

R. Cavalcante, Y. Sotomi, M. Mancone, C. Whan Lee, J. M. Ahn, Y. Onuma, P. A. Lemos, R. J. Van Geuns, S. J. Park, and P. W. Serruys. Impact of the SYNTAX scores i and II in patients with diabetes and multivessel coronary disease: A pooled analysis of patient level data from the SYNTAX, PRECOMBAT, and BEST trials. European Heart Journal, 38(25):1969–1977, 2017. ISSN 15229645.

Chassagnon, M. Vakalopoulou, A. Régent, E. I. Zacharaki, G. Aviram, C. Martin, R. Marini, N. Bus, N. Jerjir, A. Mekinian, T. Hua-Huy, L. Monnier-Cholley, N. Benmostefa, L. Mouthon, A.-T. Dinh-Xuan, N. Paragios, and M.-P. Revel. Deep Learningbased Approach for Automated Assessment of Interstitial Lung Disease in Systemic Sclerosis on CT Images. Radiology: Arti?cial Intelligence, 2(4):e190006, 2020. ISSN 2638-6100.

Cho, J. G. Lee, S. J. Kang, W. J. Kim, S. Y. Choi, J. Ko, H. S. Min, G. H. Choi, D. Y. Kang, P. H. Lee, J. M. Ahn, D. W. Park, S. W. Lee, Y. H. Kim, C. W. Lee, S. W. Park, and S. J. Park. Angiography-based machine learning for predicting fractional ?ow reserve in intermediate coronary artery lesions. Journal of the American Heart Association, 8(4):e011685, 2019. ISSN 20479980.

Couteaux, S. Si-Mohamed, R. Renard-Penna, O. Nempont, T. Lefevre, A. Popoff, G. Pizaine, N. Villain, I. Bloch, Behr, M. F. Bellin, C. Roy, O. Rouvière, S. Montagne, N. Lassau, and L. Boussel. Kidney cortex segmentation in 2D CT with U-Nets ensemble aggregation. Diagnostic and Interventional Imaging, 100(4):211–217, 2019. ISSN 22115684.

I. Cruz-Aceves, F. Oloumi, R. M. Rangayyan, J. G. Aviña-Cervantes, and A. Hernandez-Aguirre. Automatic segmentation of coronary arteries using Gabor filters and thresholding based on multiobjective optimization. Biomedical Signal Processing and Control, 25:76–85, 2016. ISSN 17468108.

T. G. Dietterich. Ensemble methods in machine learning. In Springer-Verlag Berlin Heidelberg, volume 1857 LNCS, pages 1–15, Berlin, Heidelberg, 2000. Springer Berlin Heidelberg. ISBN 3540677046.

R. El Jurdi, C. Petitjean, P. Honeine, V. Cheplygina, and F. Abdallah. A Surprisingly Effective Perimeter-based Loss for Medical Image Segmentation. In Medical Imaging with Deep Learning, pages 127–136, 2021.

H. R. Fazlali, N. Karimi, S. M. Soroushmehr, S. Shirani, B. K. Nallamothu, K. R. Ward, S. Samavi, and K. Najarian. Vessel segmentation and catheter detection in X-ray angiograms using superpixels. Medical and Biological Engineering and Computing, 56(9):1515–1530, 2018. ISSN 17410444.

M. A. Ganaie, M. Hu, M. Tanveer, and P. N. Suganthan. Ensemble deep learning: A review. arXiv preprint, page 2104.02395, 2021.

S. Gerl, J. C. Paetzold, H. He, I. Ezhov, S. Shit, F. Kofler, A. Bayat, G. Tetteh, V. Ntziachristos, and B. Menze. A Distance-Based Loss for Smooth and Continuous Skin Layer Segmentation in Optoacoustic Images. In A. L. Martel, P. Abolmaesumi, D. Stoyanov, D. Mateus, M. A. Zuluaga, S. K. Zhou, D. Racoceanu, and L. Joskowicz, editors, Springer Nature Switzerland AG, volume 12266 LNCS, pages 309–319, Cham, 2020. Springer International Publishing. ISBN 9783030597245.

D. A. Halon, D. Sapoznikov, B. S. Lewis, and M. S. Gotsman. Localization of lesions in the coronary circulation. The American Journal of Cardiology, 52(8):921–926, 1983. ISSN 00029149.

E. Hann, I. A. Popescu, Q. Zhang, R. A. Gonzales, A. Barutçu, S. Neubauer, V. M. Ferreira, and S. K. Piechnik. Deep neural network ensemble for on-the-fly quality control-driven segmentation of cardiac MRI T1 mapping. Medical Image Analysis, 71:102029, 2021. ISSN 13618423.

G. Huang, Z. Liu, L. Van Der Maaten, and K. Q. Weinberger. Densely connected convolutional networks. In Proceedings - 30th IEEE Conference on Computer Vision and Pattern Recognition, CVPR 2017, volume 2017-Janua, pages 2261–2269, 2017. ISBN 9781538604571.

Y. Huo and G. S. Kassab. The scaling of blood flow resistance: From a single vessel to the entire distal tree. Biophysical Journal, 96(2):339–346, 2009. ISSN 15420086.

I. Išgum, M. Staring, A. Rutten, M. Prokop, M. A. Viergever, and B. Van Ginneken. Multi-atlas-based segmentation with local decision fusion-application to cardiac and aortic segmentation in CT scans. IEEE Transactions on Medical Imaging, 28(7):1000–1010, 2009. ISSN 02780062.

K. Iyer, C. P. Najarian, A. A. Fattah, C. J. Arthurs, S. M. R. Soroushmehr, V. Subban, M. A. Sankardas, R. R. Nadakuditi, B. K. Nallamothu, and A. Figueroa. AngioNet: A Convolutional Neural Network for Vessel 1 Segmentation in X-ray Angiography. medRxiv, page 2021.01.25.21250488, 2021.

T. J. Jun, J. Kweon, Y. H. Kim, and D. Kim. T-Net: Nested encoderdecoder architecture for the main vessel segmentation in coronary angiography. Neural Networks, 128:216–233, 2020. ISSN 18792782.

J. Kang and J. Gwak. Ensemble of Instance Segmentation Models for Polyp Segmentation in Colonoscopy Images. IEEE Access, 7:26440–26447, 2019. ISSN 21693536.

H. Kervadec, J. Bouchtiba, C. Desrosiers, E. Granger, J. Dolz, and I. Ben Ayed. Boundary loss for highly unbalanced segmentation. Medical Image Analysis, 67:101851, 2021. ISSN 13618423.

J. Kweon, S. J. Kang, Y. H. Kim, J. G. Lee, S. Han, H. Ha, D. H. Yang, J. W. Kang, T. H. Lim, O. Kwon, J. M. Ahn, P. H. Lee, D. W. Park, S. W. Lee, C. W. Lee, S. W. Park, and S. J. Park. Impact of coronary lumen reconstruction on the estimation of endothelial shear stress: In vivo comparison of three-dimensional quantitative coronary angiography and three-dimensional fusion combining optical coherent tomography. European Heart Journal Cardiovascular Imaging, 19(10):1134–1141, 2018. ISSN 20472412.

T. C. Lee, R. L. Kashyap, and C. N. Chu. Building Skeleton Models via 3-D Medial Surface Axis Thinning Algorithms. CVGIP: Graphical Models and Image Processing, 56(6):462–478, 1994. ISSN 10499652.

D. Liang, J. Qiu, L. Wang, X. Yin, J. Xing, Z. Yang, J. Dong, J. Dong, J. Dong, and Z. Ma. Coronary angiography video segmentation method for assisting cardiovascular disease interventional treatment. BMC Medical Imaging, 20(1):65, 2020. ISSN 14712342.

F. Liu, Y. Xia, D. Yang, A. Yuille, and D. Xu. An alarm system for segmentation algorithm based on shape model. In Proceedings of the IEEE International Conference on Computer Vision, volume 2019-Octob, pages 10651–10660, 2019. ISBN 9781728148038.

M. Lorenzo-Valdés, G. I. Sanchez-Ortiz, R. Mohiaddin, and D. Rueckert. Atlas-based segmentation and tracking of 3D cardiac MR images using non-rigid registration. In T. Dohi and R. Kikinis, editors, Springer-Verlag Berlin Heidelberg, volume 2488, pages 642–650, Berlin, Heidelberg, 2002. Springer Berlin Heidelberg. ISBN 9783540457862.

Q. Lyu, H. Shan, and G. Wang. MRI Super-Resolution With Ensemble Learning and Complementary Priors. IEEE Transactions on Computational Imaging, 6:615–624, 2020. ISSN 2573-0436.

C. Mondal, M. K. Hasan, M. T. Jawad, A. Dutta, M. R. Islam, M. A. Awal, and M. Ahmad. Acute Lymphoblastic Leukemia Detection from Microscopic Images Using Weighted Ensemble of Convolutional Neural Networks. arXiv preprint, page 2105.03995, 2021.

J. H. Moon, D. Y. Lee, W. C. Cha, M. J. Chung, K. S. Lee, B. H. Cho, and J. H. Choi. Automatic stenosis recognition from coronary angiography using convolutional neural networks. Computer Methods and Programs in Biomedicine, 198:105819, 2021. ISSN 18727565.

P. D. Morris, N. Curzen, and J. P. Gunn. Angiography-derived fractional flow reserve: More or less physiology? Journal of the American Heart Association, 9(6):e015586, 2020. ISSN 20479980.

G. K. Murugesan, S. Nalawade, C. Ganesh, B. Wagner, F. F. Yu, B. Fei, A. J. Madhuranthakam, and J. A. Maldjian. Multidimensional and Multiresolution Ensemble Networks for Brain Tumor Segmentation. In A. Crimi and S. Bakas, editors, Springer Nature Switzerland AG, volume 12658 LNCS, pages 448–457, Cham, 2021. Springer International Publishing. ISBN 9783030720834.

M. Ono, P. W. Serruys, M. R. Patel, J. Escaned, T. Akasaka, M. A. Lavieren, C. Haase, M. Grass, N. Kogame, H. Hara, H. Kawashima, J. J. Wykrzykowska, J. J. Piek, S. Garg, N. O’Leary, B. Inderbitzen, and Y. Onuma. A prospective multicenter validation study for a novel angiography-derived physiological assessment software: Rationale and design of the radiographic imaging validation and evaluation for Angio-iFR (ReVEAL iFR) study. American Heart Journal, 239:19–26, 2021. ISSN 10976744.

B. Qin, M. Jin, D. Hao, Y. Lv, Q. Liu, Y. Zhu, S. Ding, J. Zhao, and B. Fei. Accurate vessel extraction via tensor completion of background layer in X-ray coronary angiograms. Pattern Recognition, 87:38–54, 2019. ISSN 00313203.

V. Sandfort, K. Yan, P. M. Graffy, P. J. Pickhardt, and R. M. Summers. Use of Variational Autoencoders with Unsupervised Learning to Detect Incorrect Organ Segmentations at CT. Radiology: Artificial Intelligence, 3(4):e200218, 2021. ISSN 2638-6100.

T. Schlegl, P. Seeböck, S. M. Waldstein, U. Schmidt-Erfurth, and G. Langs. Unsupervised anomaly detection with generative adversarial networks to guide marker discovery. In M. Niethammer, M. Styner, S. Aylward, H. Zhu, I. Oguz, P.-T. Yap, and D. Shen, editors, LSpringer Nature Switzerland AG, volume 10265 LNCS, pages 146–147, Cham, 2017. Springer International Publishing. ISBN 9783319590493.

R. Shah, E. Yow, W. S. Jones, L. P. Kohl, A. S. Kosinski, U. Hoffmann, K. L. Lee, C. B. Fordyce, D. B. Mark, A. Lowe, P. S. Douglas, and M. R. Patel. Comparison of visual assessment of coronary stenosis with independent quantitative coronary angiography: Findings from the Prospective Multicenter Imaging Study for Evaluation of Chest Pain (PROMISE) trial. American Heart Journal, 184:1–9, 2017. ISSN 10976744.

S. Shit, J. C. Paetzold, A. Sekuboyina, I. Ezhov, A. Unger, A. Zhylka, J. P. W. Pluim, U. Bauer, and B. H. Menze. Cldice - a novel topology-preserving loss function for tubular structure segmentation. In Proceedings of the IEEE/CVF Conference on Computer Vision and Pattern Recognition (CVPR), pages 16560–16569, June 2021.

G. Sianos, M.-A. Morel, A. P. Kappetein, M.-C. Morice, A. Colombo, K. Dawkins, M. van den Brand, N. Van Dyck, M. E. Russell, F. W. Mohr, and P. W. Serruys. The SYNTAX Score: an angiographic tool grading the complexity of coronary artery disease. EuroIntervention : journal of EuroPCR in collaboration with the Working Group on Interventional Cardiology of the European Society of Cardiology, 1(2):219–27, 2005. ISSN 1774-024X.

J. L. Silva, M. N. Menezes, T. Rodrigues, B. Silva, F. J. Pinto, and A. L. Oliveira. Encoder-Decoder Architectures for Clinically Relevant Coronary Artery Segmentation. arXiv preprint, page 2106.11447, 2021.

A. Sohail, A. Khan, H. Nisar, S. Tabassum, and A. Zameer. Mitotic nuclei analysis in breast cancer histopathology images using deep ensemble classifier. Medical Image Analysis, 72:102121, 2021. ISSN 13618423.

P. H. Stone, S. Saito, S. Takahashi, Y. Makita, S. Nakamura, T. Kawasaki, A. Takahashi, T. Katsuki, S. Nakamura, A. Namiki, A. Hirohata, T. Matsumura, S. Yamazaki, H. Yokoi, S. Tanaka, S. Otsuji, F. Yoshimachi, J. Honye, D. Harwood, M. Reitman, A. U. Coskun, M. I. Papafaklis, and C. L. Feldman. Prediction of progression of coronary artery disease and clinical outcomes using vascular profiling of endothelial shear stress and arterial plaque characteristics: The PREDICTION study. Circulation, 126(2):172–181, 2012. ISSN 00097322.

V. Sundaresan, G. Zamboni, P. M. Rothwell, M. Jenkinson, and L. Griffanti. Triplanar ensemble U-Net model for white matter hyperintensities segmentation on MR images. Medical Image Analysis, 73:102184, 2021. ISSN 13618423.

D. J. Thuijs, A. P. Kappetein, P. W. Serruys, F. W. Mohr, M. C. Morice, M. J. Mack, D. R. Holmes, N. Curzen, P. Davierwala, T. Noack, M. Milojevic, K. D. Dawkins, B. R. da Costa, P. Jüni, S. J. Head, F. Casselman, B. de Bruyne, E. Høj Christiansen, J. M. Ruiz-Nodar, P. Vermeersch, W. Schultz, M. Sabaté, G. Guagliumi, H. Grubitzsch, K. Stangl, O. Darremont, M. Bentala, P. den Heijer, I. Preda, R. Stoler, T. Szerafin, J. K. Buckner, M. S. Guber, N. Verberkmoes, F. Akca, T. Feldman, F. Beyersdorf, B. Drieghe, K. Oldroyd, G. Berg, A. Jeppsson, K. Barber, K. Wolschleger, J. Heiser, P. van der Harst, M. A. Mariani, H. Reichenspurner, C. Stark, M. Laine, P. C. Ho, J. C. Chen, R. Zelman, P. A. Horwitz, A. Bochenek, A. Krauze, C. Grothusen, D. Dudek, G. Heyrich, P. Kolh, V. LeGrand, P. Coelho, S. Ensminger, B. Nasseri, R. Ingemansson, G. Olivecrona, J. Escaned, R. Guera, S. Berti, A. Chieffo, N. Burke, M. Mooney, A. Spolaor, C. Hagl, M. Näbauer, M. J. Suttorp, R. A. Stine, T. McGarry, S. Lucas, K. Endresen, A. Taussig, K. Accola, U. Canosi, I. Horvath, L. Cannon, J. D. Talbott, C. W. Akins, R. Kramer, M. Aschermann, W. Killinger, I. Narbute, F. Burzotta, A. Bogers, F. Zijlstra, H. Eltchaninoff, J. Berland, G. Stefanini, I. Cruz Gonzalez, U. Hoppe, S. Kiesz, B. Gora, A. Ahlsson, M. Corbascio, T. Bilfinger, D. Carrie, D. Tchétché, K. E. Hauptman, E. Stahle, S. James, S. Sandner, G. Laufer, I. Lang, A. Witkowski, V. Thourani, H. Suryapranata, S. Redwood, C. Knight, P. MacCarthy, A. de Belder, A. Banning, and A. Gershlick. Percutaneous coronary intervention versus coronary artery bypass grafting in patients with three-vessel or left main coronary artery disease: 10-year followup of the multicentre randomised controlled SYNTAX trial. The Lancet, 394(10206):1325–1334, 2019. ISSN 1474547X.

M. Tomaniak, Y. Katagiri, R. Modolo, R. de Silva, R. Y. Khamis, C. V. Bourantas, R. Torii, J. J. Wentzel, F. J. Gijsen, G. van Soest, P. H. Stone, N. E. West, A. Maehara, A. Lerman, A. F. van der Steen, T. F. Lüscher, R. Virmani, W. Koenig, G. W. Stone, J. E. Muller, W. Wijns, P. W. Serruys, and Y. Onuma. Vulnerable plaques and patients: State-of-the-art. European Heart Journal, 41(31):2997–3004, 2020. ISSN 15229645.

S. Tu, E. Barbato, Z. Köszegi, J. Yang, Z. Sun, N. R. Holm, B. Tar, Y. Li, D. Rusinaru, W. Wijns, and J. H. Reiber. Fractional flow reserve calculation from 3-dimensional quantitative coronary angiography and TIMI frame count: A fast computer model to quantify the functional significance of moderately obstructed coronary arteries. JACC: Cardiovascular Interventions, 7(7):768–777, 2014. ISSN 18767605.

T. Wan, J. Chen, Z. Zhang, D. Li, and Z. Qin. Automatic vessel segmentation in X-ray angiogram using spatiotemporal fully-convolutional neural network. Biomedical Signal Processing and Control, 68:102646, 2021. ISSN 17468108.

L. Wang, D. Liang, X. Yin, J. Qiu, Z. Yang, J. Xing, J. Dong, and Z. Ma. Coronary artery segmentation in angiographic videos utilizing spatial-temporal information. BMC medical imaging, 20(1):110, 2020. ISSN 14712342.

A. J. Weisman, M. W. Kieler, S. B. Perlman, M. Hutchings, R. Jeraj, L. Kostakoglu, and T. J. Bradshaw. Convolutional Neural Networks for Automated PET/CT Detection of Diseased Lymph Node Burden in Patients with Lymphoma. Radiology: Artificial Intelligence, 2(5):e200016, 2020. ISSN 2638-6100.

S. Winzeck, S. J. Mocking, R. Bezerra, M. J. Bouts, E. C. Mcintosh, I. Diwan, P. Garg, A. Chutinet, W. T. Kimberly, W. A. Copen, P. W. Schaefer, H. Ay, A. B. Singhal, K. Kamnitsas, B. Glocker, A. G. Sorensen, and O. Wu. Ensemble of convolutional neural networks improves automated segmentation of acute ischemic lesions using multiparametric diffusion-weighted MRI. American Journal of Neuroradiology, 40(6):938–945, 2019. ISSN 1936959X.

W. Wu, S. Samant, G. de Zwart, S. Zhao, B. Khan, M. Ahmad, M. Bologna, Y. Watanabe, Y. Murasato, F. Burzotta, E. S. Brilakis, G. Dangas, Y. Louvard, G. Stankovic, G. S. Kassab, F. Migliavacca, C. Chiastra, and Y. S. Chatzizisis. 3D reconstruction of coronary artery bifurcations from coronary angiography and optical coherence tomography: feasibility, validation, and reproducibility. Scientific Reports, 10(1):18049, 2020. ISSN 20452322.

Y. Xia, Y. Zhang, F. Liu, W. Shen, and A. L. Yuille. Synthesize Then Compare: Detecting Failures and Anomalies for Semantic Segmentation. In A. Vedaldi, H. Bischof, T. Brox, and J.-M. Frahm, editors, Springer Nature Switzerland AG, volume 12346 LNCS, pages 145–161, Cham, 2020. Springer International Publishing. ISBN 9783030584511.

S. Yang, J. Kweon, J. H. Roh, J. H. Lee, H. Kang, L. J. Park, D. J. Kim, H. Yang, J. Hur, D. Y. Kang, P. H. Lee, J. M. Ahn, S. J. Kang, D. W. Park, S. W. Lee, Y. H. Kim, C. W. Lee, S. W. Park, and S. J. Park. Deep learning segmentation of major vessels in X-ray coronary angiography. Scientific Reports, 9(1):16897, 2019. ISSN 20452322.

M. Zhai, T. Du, R. Yang, and H. Zhang. Coronary Artery Vascular Segmentation on Limited Data via Pseudo-Precise Label. In Proceedings of the Annual International Conference of the IEEE Engineering in Medicine and Biology Society, EMBS, pages 816–819, 2019. ISBN 9781538613115.

S. Zhai, R. Gu, W. Lei, and G. Wang. Myocardial Edema and Scar Segmentation Using a Coarse-to-Fine Framework with Weighted Ensemble. In X. Zhuang and L. Li, editors, Springer Nature Switzerland AG, volume 12554 LNCS, pages 49–59, Cham, 2020. Springer International Publishing. ISBN 9783030656508.

D. Zhang, G. Yang, S. Zhao, Y. Zhang, D. Ghista, H. Zhang, and S. Li. Direct Quantification of Coronary Artery Stenosis through Hierarchical Attentive Multi-View Learning. IEEE Transactions on Medical Imaging, 39(12): 4322–4334, 2020. ISSN 1558254X.

C. Zhao, A. Vij, S. Malhotra, J. Tang, H. Tang, D. Pienta, Z. Xu, and W. Zhou. Automatic extraction and stenosis evaluation of coronary arteries in invasive coronary angiograms. Computers in Biology and Medicine, 136:104667, 2021. ISSN 18790534.

L. Zhao, D. Li, J. Chen, and T. Wan. Automated Coronary Tree Segmentation for X-ray Angiography Sequences Using Fully-convolutional Neural Networks. In VCIP 2018 - IEEE International Conference on Visual Communications and Image Processing, pages 1–4, 2018. ISBN 9781538644584.

H. Zheng, Y. Zhang, L. Yang, P. Liang, Z. Zhao, C. Wang, and D. Z. Chen. A new ensemble learning framework for 3D biomedical image segmentation. In 33rd AAAI Conference on Artificial Intelligence, AAAI 2019, 31st Innovative Applications of Artificial Intelligence Conference, IAAI 2019 and the 9th AAAI Symposium on Educational Advances in Artificial Intelligence, EAAI 2019, volume 33, pages 5909–5916, 2019. ISBN 9781577358091.

Z. H. Zhou, J. Wu, and W. Tang. Ensembling neural networks: Many could be better than all. Artificial Intelligence, 137(1-2):239–263, 2002. ISSN 00043702.

X. Zhu, Z. Cheng, S. Wang, X. Chen, and G. Lu. Coronary angiography image segmentation based on PSPNet. Computer Methods and Programs in Biomedicine, 200:105897, 2021. ISSN 18727565.

